# Most coronial recommendations in Australia have no published response: a national analysis of findings and agency responses

**DOI:** 10.64898/2026.02.12.26346201

**Authors:** Hayden Farquhar

## Abstract

**Background:** A coroner’s recommendation prevents deaths only if someone acts on it. We measured how often Australian coronial recommendations receive a published response and what it says.

**Methods:** Cross-sectional analysis of every finding and agency response in the AustLII Coronial Law Library (eight jurisdictions; enumerated February 2026). Responses were linked to findings and classified by a large language model, validated against the author’s blinded coding (seven-category kappa 0.81, 95% CI 0.71 to 0.90). Response rates were measured over all years and within each jurisdiction’s response-database years. Acceptance was modelled by Firth penalised logistic regression.

**Results:** Of 9,831 findings, 4,480 (45.6%) contained recommendations. Only 1,309 of these (29.2%) had a published agency response. Within the database years the figure was 36.6% (35.0% to 38.3%), from 58.6% in Western Australia to 5.8% in Tasmania; the Australian Capital Territory has no response database. Of 1,541 findings with a classifiable response, 1,316 (85.4%; 83.5% to 87.1%) accepted the recommendations, from 65.0% in Western Australia to all 22 in the Northern Territory. Findings that mentioned a psychiatric unit (adjusted odds ratio (OR) 2.02, 1.25 to 3.35) or a prison (1.78, 1.12 to 2.92) were more likely to receive an accepting response. A mention of Aboriginal or Torres Strait Islander identity was not (0.78, 0.46 to 1.37).

**Conclusions:** Most coronial recommendations in Australia have no response that anyone can find, and those that are answered in public are mostly accepted. A national register linking each recommendation to its response would make implementation, not only stated intent, possible to follow up.

## Introduction

Coroners are among the few officials whose job includes saying how the next death could be prevented. In every Australian jurisdiction they may recommend changes to policy, practice or law (table 2). Studies of Victorian recommendations and of river drowning found that most specify a countermeasure and the level at which it should act, though many rely on administrative controls rather than stronger measures^1,2^, and recent work has mapped the themes of recommendations to guide prevention^3^. A recommendation prevents nothing until someone acts on it, however, and evidence on what happens next is thin. In England and Wales, where recipients of a Prevention of Future Deaths report must reply, topic-specific case series find a published response for only 40% to 78% of reports or report recipients^4-11^.

What happens to a recommendation once it leaves the court depends on where it was made (table 2). Four jurisdictions impose a statutory duty to respond. In Victoria a public entity must reply within three months with “a statement of action (if any) that has, is or will be taken”, and the coroner must publish the reply online^12^. South Australia, the Australian Capital Territory and the Northern Territory route responses to Parliament, the Legislative Assembly or the Attorney-General^13-15^.

Queensland’s Act requires the coroner to send comments to government and publish them, but not that anyone reply^16^. Its published responses rest on an administrative arrangement introduced in 2008^17,18^. New South Wales relies on a Premier’s memorandum asking agencies to report within six months^19^, and Western Australia and Tasmania have no response requirement^20,21^. The Victorian and Queensland Acts were amended in 2026, after the study period, without changing these provisions.

Australian evidence on responses comes from single jurisdictions or single death types. In Victoria’s first years under s 72, recipient organisations reported implementing 37% of recommendations and rejecting 27%^22^, and only 44% of written responses said explicitly whether action had been or would be taken^23^. A national study of recommendations after the homicide of older adults found a response for 27 of 29 recommendations, most of them supported^24^. Recent work has applied language models to national collections of coronial reports, but to the circumstances of death rather than to responses^25,26^. No study has measured, across all jurisdictions and causes of death, how often a recommendation receives a published response and what that response says.

We used the national open collection of coronial findings and responses to estimate, for each Australian jurisdiction, the proportion of findings with recommendations that have a published agency response and the proportion of responses that accept the recommendations, and to examine which characteristics of the death are associated with acceptance.

## Methods

### Data sources

We enumerated every document in the coronial databases of the Australasian Legal Information Institute (AustLII) Coronial Law Library in February 2026^27,28^, which hold findings from all eight Australian jurisdictions and agency responses from seven (the Australian Capital Territory has no response database). The findings databases held 9,924 records, two of them duplicates. Of the 9,922 findings, 9,831 had usable text (37 were under 200 characters and 54 were empty). The response databases held 2,064 documents.

Text was extracted from HTML and PDF documents, with optical character recognition for scanned pages. An audit of every PDF found that the original extraction had read only the text layer of mixed documents; 741 PDFs (158 findings and 583 responses) were re-extracted page by page. Most of the affected responses were Victorian: AustLII stores many Victorian responses as an index page followed by scanned agency letters. For 473 such records the letters were already on disk; 110 records held only the index page, and we retrieved the listed responses from the Coroners Court of Victoria website and the Internet Archive, accepting a document only when its text named the deceased. Responses were retrieved for 100 of the 110.

AustLII publishes what each court supplies, so the corpus is the public record of the coronial system rather than all reportable deaths, and we expect it to be enriched for inquests and for cases with recommendations.

### Linking responses to findings

Responses were linked to findings by court reference numbers where available (Victoria, Queensland, Tasmania and the Northern Territory) and otherwise by the deceased’s name and year (New South Wales, Western Australia and South Australia). Documents without a usable name were not name-matched (table S1).

Each response document was assigned a status (table S2). Agency responses counted, as did 20 documents showing that a response existed but whose text was unavailable (including the 10 outstanding Victorian records). Court tables recording that a response was awaited, not required or not received did not count. There are two co-primary response measures: the proportion of findings with recommendations that have at least one counting response over all years, and the same proportion within each jurisdiction’s response-database years (from the first to the last year in which the database holds a counting response) for findings up to 2023, so that every finding had at least two years to be answered. The second was added after the first results were seen, when it became clear that the databases cover different periods (table 1).

**Table 1.** Findings, recommendations, published responses and acceptance by jurisdiction.

| Jurisdiction | Findings | Years | With recommendations, n (%) | Published response, all years, n (%) | Response database years | Published response within database years, n/N (%) | Classifiable findings | Accepted, n (%; 95% CI) |
|---|---|---|---|---|---|---|---|---|
| NSW | 1,071 | 1979-2026 | 435 (40.6) | 206 (47.4) | 2008-2025 | 172/343 (50.1) | 290 | 273 (94.1; 90.8 to 96.3) |
| Vic | 4,324 | 2002-2025 | 1,664 (38.5) | 700 (42.1) | 2009-2021 | 700/1,399 (50.0) | 774 | 633 (81.8; 78.9 to 84.3) |
| Qld | 1,035 | 2004-2026 | 565 (54.6) | 161 (28.5) | 2008-2025 | 158/450 (35.1) | 202 | 185 (91.6; 86.9 to 94.7) |
| WA | 705 | 2012-2025 | 204 (28.9) | 111 (54.4) | 2013-2025 | 99/169 (58.6) | 120 | 78 (65.0; 56.1 to 72.9) |
| SA | 884 | 2000-2025 | 620 (70.1) | 73 (11.8) | 2005-2024 | 69/454 (15.2) | 79 | 75 (94.9; 87.7 to 98.0) |
| Tas | 1,304 | 2001-2026 | 733 (56.2) | 37 (5.0) | 2015-2025 | 32/549 (5.8) | 54 | 50 (92.6; 82.4 to 97.1) |
| ACT | 96 | 2013-2025 | 42 (43.8) | no response database |  |  |  |  |
| NT | 412 | 1981-2026 | 217 (52.7) | 21 (9.7) | 2019-2025 | 12/28 (42.9) | 22 | 22 (100.0; 85.1 to 100.0) |
| Total | 9,831 |  | 4,480 (45.6) | 1,309 (29.2) |  | 1,242/3,392 (36.6) | 1,541 | 1,316 (85.4; 83.5 to 87.1) |

Years are AustLII publication years. Response database years: first and last publication year of a counting response; the within-database rate covers findings from those years up to 2023 (Australian Capital Territory excluded from the total). Classifiable findings are findings with at least one counting response whose position could be classified; they include 244 findings with a response in which the recommendation detector found no recommendation, so in some jurisdictions they exceed the response column. Accepted: implemented, already implemented or partially accepted.

### Classifying responses

Each response was classified by a large language model (Claude Opus 5.5, Anthropic) using a prompt written from the author’s coding definitions and fixed before testing, under a dated protocol written before each step and deposited with the analysis code^29^. The model assigned the document type and one of seven positions: implemented, already implemented, partially accepted, under consideration, noted, not supported, or unclassifiable. Accepted means implemented, already implemented or partially accepted. These categories record the position stated in the response, not whether anything changed in practice.

The prompt and model were tested once against 100 responses that the author had coded blind to any machine label. Seven-category agreement was kappa 0.81 (95% CI 0.71 to 0.90) and binary acceptance agreement 0.73 (0.50 to 0.91), against 0.17 and 0.10 for a rule-based classifier tested first (tables S3 and S4). Because only 10 of the 90 classifiable test responses were not accepted, the author coded 40 more from the full corpus, blind to the model’s labels: 30 the model labelled not accepted and 10 accepted. The author agreed in 25 of 30 (95% CI 65% to 94%) and 10 of 10 (69% to 100%); weighted to the corpus, binary accuracy was 97.2% (table S5).

Where a finding had more than one counting response, its outcome was the position taken by most of its classifiable responses, with an exact tie counted as partially accepted (the rule the author’s coding definitions apply to a single letter answering several recommendations). This rule was fixed before the models were refitted; accepted if any response accepts, and accepted only if all do, are reported as sensitivity analyses (table S6).

### Case characteristics

Indigenous status, medication involvement and custodial or institutional settings (psychiatric facility, prison or other correctional setting, police custody) were flagged by layered regular expressions on the text of the findings (table S7). These flags mark what a finding mentions, not the place of death or a coded attribute of the deceased. A post hoc sensitivity analysis refitted the acceptance model with narrower flags that excluded community crisis teams, police watch houses and remand.

### Statistical analysis

Proportions are given with Wilson 95% confidence intervals. Whether a response accepted the recommendations was modelled by logistic regression on jurisdiction (reference New South Wales), year, log document length, and the case flags. Because every Northern Territory response was accepted, the models used Firth’s penalised likelihood with profile-likelihood confidence intervals^30,31^; an ordinary model excluding the Northern Territory is reported as a sensitivity analysis. Benjamini-Hochberg correction^32^ was applied across all 41 tests run in the analysis, including exploratory tests of recommendation presence and document length that are not reported here (table S8). Analyses used Python 3.14 and statsmodels 0.15; the analysis code and derived data are public^29^.

### Ethics and Indigenous data

All documents are court records already published by the courts. The study involved no contact with families and reports no names or case details, so formal ethics review was not sought and consent was not applicable. The documents remain subject to the terms of AustLII and the courts and are not redistributed. Some findings concern the deaths of Aboriginal and Torres Strait Islander people. No Aboriginal or Torres Strait Islander organisation or community was involved in the design or interpretation of this analysis, which falls short of the collective benefit and authority to control set out in the CARE and Maiam nayri Wingara principles^33,34^. Results are therefore reported only in aggregate, and the analysis draws no conclusions about Aboriginal and Torres Strait Islander communities.

## Results

### Recommendations and published responses

Of 9,831 findings, 4,480 (45.6%; 95% CI 44.6% to 46.6%) contained recommendations, ranging from 28.9% in Western Australia to 70.1% in South Australia (table 1).

Only 1,309 of the 4,480 findings with recommendations (29.2%) had a counting response in the AustLII record. The proportion was highest in Western Australia (54.4%), New South Wales (47.4%) and Victoria (42.1%), and lowest in South Australia (11.8%), the Northern Territory (9.7%), Tasmania (5.0%) and the Australian Capital Territory, which has no response database. In New South Wales a further 31 findings (7.1% of those with recommendations) had only a court table recording that a response was awaited; counting any linked document, the New South Wales figure would be 55.2% (table S9).

The response databases cover different years (table 1 and table S10). Victoria’s holds responses published from 2009 to 2021, Tasmania’s from 2015 and the Northern Territory’s from 2019. Within each jurisdiction’s database years, 1,242 of 3,392 findings with recommendations (36.6%; 35.0% to 38.3%) had a counting response. The order changed little except in the Northern Territory, where the rate rose from 9.7% to 42.9% (12 of 28). South Australia (15.2%) and Tasmania (5.8%) remained low (figure 1).

**Figure 1.**
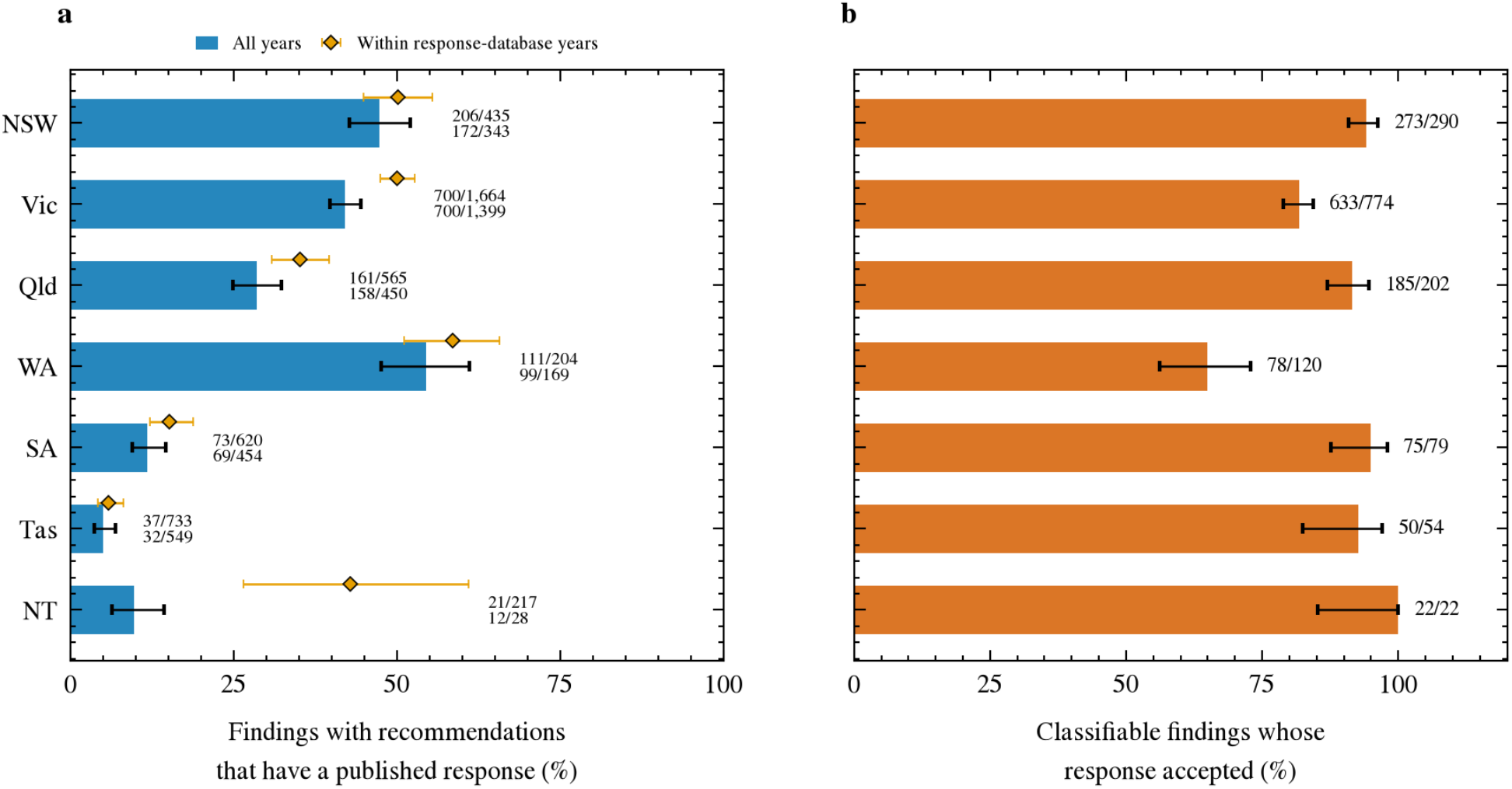
Published responses and acceptance by jurisdiction. (a) Findings with recommendations that have at least one published agency response on AustLII, over all years (bars) and within each jurisdiction’s response-database years up to 2023 (diamonds); labels give counts for each. (b) Classifiable findings whose response accepted the recommendations (implemented, already implemented or partially accepted). Bars and diamonds show percentages with Wilson 95% confidence intervals. The Australian Capital Territory has no response database and is omitted.

### What responses said

In all, 1,541 findings had a counting response with a classifiable outcome. They include 244 findings in which the recommendation detector had found no recommendation, and acceptance was the same with or without them (85.4%). Of the 1,541, 1,316 (85.4%; 83.5% to 87.1%) accepted the recommendations: 64.0% were implemented, 6.9% already implemented and 14.5% partially accepted. A further 8.3% were under consideration, 5.0% not supported and 1.3% noted. Acceptance ranged from 65.0% (56.1% to 72.9%) in Western Australia, where many responses left recommendations under consideration, to 100% of the 22 Northern Territory findings (table 1).

### Case characteristics and acceptance

In the adjusted model (figure 2), acceptance was lower in Victoria (OR 0.29, 95% CI 0.16 to 0.50) and Western Australia (0.12, 0.06 to 0.21) than in New South Wales. Findings that mentioned a psychiatric unit (2.02, 1.25 to 3.35) or a prison or other correctional setting (1.78, 1.12 to 2.92) were more likely to receive an accepting response. A mention of Aboriginal or Torres Strait Islander identity was not associated with acceptance (0.78, 0.46 to 1.37), nor was year (0.98 per year, 0.94 to 1.03). Both facility associations survived false discovery rate correction, along with 27 other tests (29 of 41 in all).

**Figure 2.**
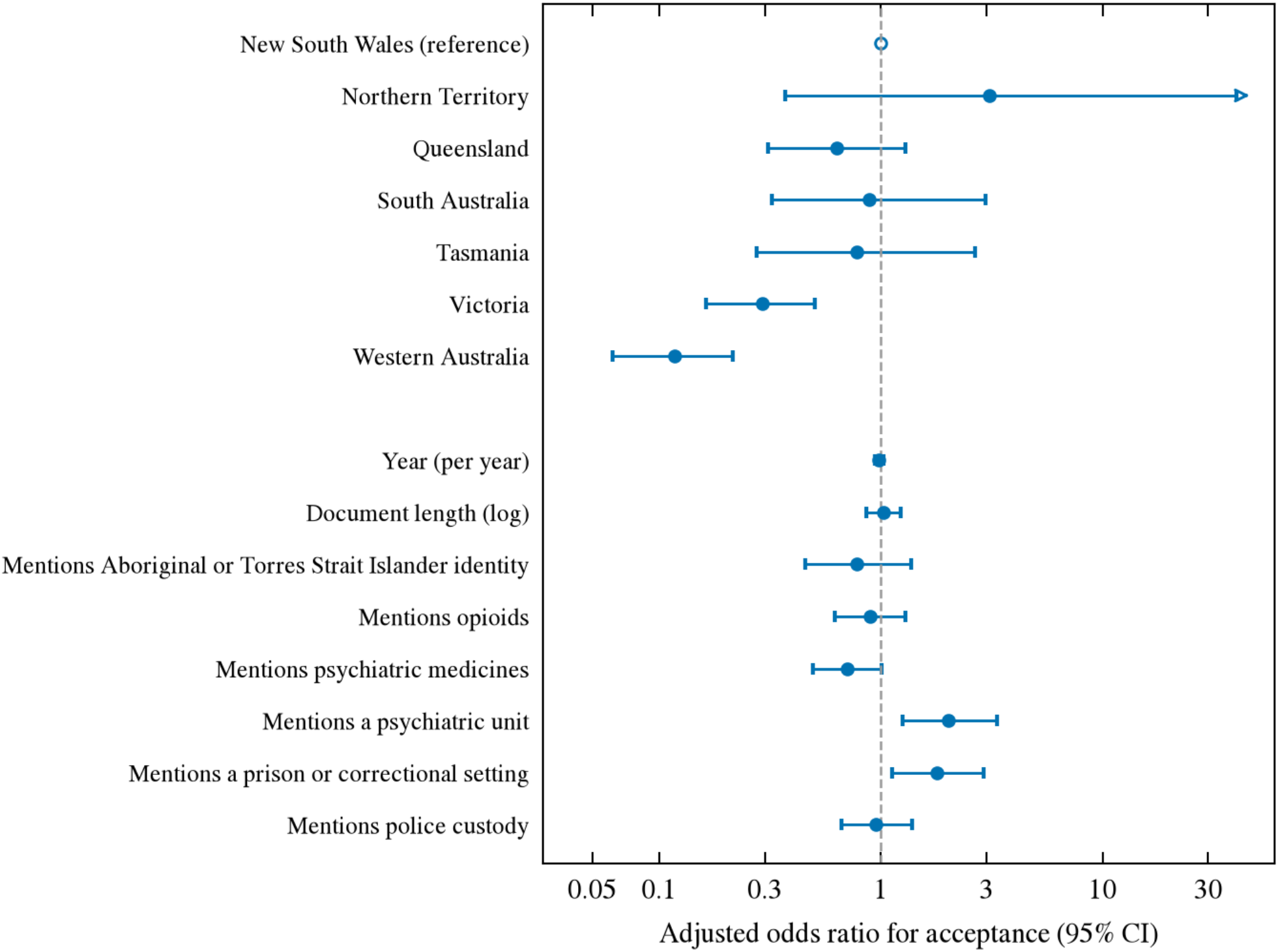
Adjusted odds ratios for an accepting response (n = 1,541 findings), from a Firth penalised logistic regression. Reference jurisdiction: New South Wales. The Northern Territory upper limit (402) is truncated (arrow). Case characteristics are flags for what a finding mentions (table S7).

The facility associations held under the alternative rules for findings with several responses (psychiatric facility 2.11 if any response accepted and 1.61 if all had to; prison 1.83 and 1.59) and with the narrower flags (post hoc: 1.81, 1.11 to 3.05; and 1.71, 1.07 to 2.82). Excluding the Northern Territory changed no other odds ratio by more than 0.08. The jurisdiction contrasts were sensitive to the reference group, which had only 17 non-accepting findings: under the strictest rule, the Victorian odds ratio was 0.51 (0.31 to 0.82).

## Discussion

Across Australia, fewer than a third of coronial findings with recommendations had a published agency response on AustLII, and little more than a third even within the years the response databases cover. When a response was published, it usually accepted the recommendations: 85% of findings with a classifiable response were implemented, already implemented or partially accepted. The weak point in this record is therefore less what agencies say than whether anyone can see that they said it.

### Visibility depends on where responses go, not only on whether they are required

The proportion of findings with a published response did not line up with the statutory duties in table 2. Within the databases’ years, Victoria (a duty to respond and to publish) and New South Wales and Western Australia (no statutory duty) all had published responses for half or more of findings with recommendations. South Australia, which requires ministers to report to Parliament, reached 15%, and Tasmania, with no requirement, 6%. Most Northern Territory findings with recommendations (164 of 217) predate its response database, which begins in 2019; within that period 12 of 28 had a response. The Australian Capital Territory, despite its tabling duty, has no response database at all. A duty to respond and a place where responses can be found are separate design choices, and only the second lets families, researchers and other agencies learn what happened to a recommendation. The gap also shows within jurisdictions. Victoria’s Act requires online publication, yet no Victorian response after 2021 is on AustLII, and Queensland’s court publishes responses beside its findings^18^ while only 28.5% of Queensland findings with recommendations have one on AustLII. The figures here are a floor for what exists and an accurate count of what the national collection holds.

**Table 2.** Arrangements for responding to coronial recommendations, by jurisdiction.

| **Jurisdiction** | **Statutory duty to respond** | **Who responds, to whom, and when** | **Publication of responses** |
|---|---|---|---|
| NSW | No | Premier’s Memorandum M2009-12: agencies acknowledge within 21 days and report to the Attorney General within 6 months, with reasons if not implementing | Attorney General’s department reports twice a year (M2009-12); responses appear on AustLII |
| Vic | Yes, s 72(3)-(4) | Public statutory authority or entity, to the coroner, within 3 months; must state the action (if any) taken or to be taken | Published online by the coroner (s 72(5)) |
| Qld | No | Administrative arrangement since 2008: report on each recommendation within 6 months, updated twice a year | Published next to the findings since 2013 |
| WA | No | No requirement | Responses appear on AustLII |
| SA | Yes, s 25(5) | Each relevant Minister, to each House of Parliament, within 8 sitting days after 6 months; action taken or proposed, or reasons for none | Laid before Parliament |
| Tas | No | No requirement | None located |
| ACT | Yes, ss 57(4), 76 | Attorney-General or Minister presents the report and a response to the Legislative Assembly; for deaths in care or custody, the agency responds to its Minister within 3 months | Presented to the Legislative Assembly |
| NT | Yes, s 46B | Agency chief executive or Commissioner of Police, to the Attorney-General, within 3 months; Attorney-General tables a report | Tabled in the Legislative Assembly |

Sources: the jurisdictions’ Coroners Acts, Queensland State Coroner’s Guidelines chapter 9 and NSW Premier’s Memorandum M2009-12<sup>12-21,27</sup>. Law as in force during the study period. Changes during the period: South Australia substituted s 25(2) and (5) and inserted s 25(6) to (8) from 7 June 2021 (Coroners (Inquests and Privilege) Amendment Act 2021), and the Australian Capital Territory amended s 57 in 2019 and s 76 from 29 January 2021 (Coroners Amendment Act 2020). Changes after the period, in 2026: Victoria’s Justice Legislation Further Amendment (Miscellaneous) Act 2026 added powers to discontinue some investigations and changed who may apply to set findings aside, and Queensland’s Justice and Other Legislation Amendment Act 2026 amended the provisions on findings and comments (ss 45 to 47, from 29 October 2026); neither altered the response arrangements shown.

The same pattern appears in England and Wales, where responses are mandatory and yet case series across topics find a published response for 40% to 78% of reports or report recipients^4-11^; larger organisations respond more often than smaller ones^35^, and many responses never enter the public domain^36^. A response that exists only in a departmental file cannot inform prevention elsewhere.

### What responses said

Stated acceptance was high and fairly uniform, with two exceptions. In Western Australia a third of findings with a response were not accepted, mostly because the response left the recommendation under consideration, possibly because responses there are published before a decision is made.

Victorian responses accepted less often than those in New South Wales (81.8% against 94.1%). One explanation, which these data cannot test, is selection: where responding is voluntary, the replies that are written and published may lean towards acceptance, while a mandatory regime also captures those that decline or merely note a recommendation. The contrast narrowed (odds ratio 0.51) when only findings whose every response accepted counted as accepted.

High stated acceptance is not the same as implementation. Victorian organisations surveyed directly reported implementing 37% of recommendations^22^, and fewer than half of Victorian responses stated plainly what would be done^23^; in the older-adult homicide study, most recommendations were supported, and over half had been implemented or partly implemented when the study was done^24^. A government review reported most recommendations of the Royal Commission into Aboriginal Deaths in Custody implemented^37^, a conclusion disputed using later coronial findings^38^. Our measure records the position an agency took on paper, and should be read as an upper bound on change in practice.

### Psychiatric and custodial settings

Findings that mentioned a psychiatric unit or a prison were more likely to receive an accepting response, after adjustment, under each rule for findings with several responses and with narrower flags. The flags record a mention, not necessarily the place of death, but these are settings run or closely regulated by government, where the agency that receives a recommendation is usually the one able to act on it and deaths attract sustained scrutiny. A mention of Aboriginal or Torres Strait Islander identity, which the flag detects through identity terms, organisations or context such as remote community or customary law, was not associated with acceptance once jurisdiction was taken into account. The null result says nothing about whether accepted recommendations were implemented.

### Strengths and limitations

This is the first analysis of published responses across all Australian jurisdictions and causes of death, with a classifier fixed before testing and validated twice against blinded human coding. AustLII holds what courts supply and keeps adding documents, including some dated before the enumeration, so findings are enriched for inquests and the response measures count published responses, not all responses. In New South Wales, where linkage relied on names, 57% of response documents could be linked and the links were not checked by hand; of the 291 unlinked, 207 named a person with no New South Wales finding on AustLII, 57 had no usable name, and only 22 may be missed links (table S1).

Ten Victorian and four New South Wales responses known to exist were unavailable; they count as responses but not towards acceptance. The classifier’s errors in the blinded check ran one way, so acceptance is if anything slightly understated. Case characteristics came from text, not coded data. The within-database measure was defined after the first results were seen and uses the years each database holds responses as a proxy for when they were collected. Finally, the jurisdiction contrasts rest on a reference group with few non-accepting findings.

### Implications

Australia lacks a single place where a recommendation and its response can be read together. A national register, built on Victorian and Queensland publication practice, would make the response rate visible and allow follow-up of implementation, not only of stated intent. The methods used here can be rerun as findings are published, so such a register could report response and acceptance rates routinely.

## Conclusion

In Australia a coronial recommendation is more likely to go unanswered in public than to be refused. Making responses findable, and then checking what happened, would do more for prevention than more evidence that agencies agree.

## Data Availability

The underlying coronial findings are publicly available on the Australasian Legal Information Institute (AustLII) Coronial Law Library (https://www.austlii.edu.au/au/special/coronial/). AustLII's terms of use do not permit bulk redistribution of source documents; however, the raw corpus can be reconstructed using the scraping scripts provided in the companion repository. All analysis code, derived output tables (including topic assignments, compliance classifications, and summary statistics), the trained recommendation topic model, and all figures are available at https://github.com/hayden-farquhar/Coronial-NLP-Analysis. No individual-level data beyond the publicly available court records were used.

## Declarations

### Patient and public involvement

Patients and the public were not involved in the design, conduct, reporting or dissemination of this research.

### Ethics approval

Not sought; the study used only court records already published by the courts (see Methods).

### Data availability statement

All source documents are publicly available from AustLII. The analysis code, the frozen classifier prompt and protocol, the classifier labels and the derived finding-level dataset are available at https://github.com/hayden-farquhar/Coronial-NLP-Analysis and archived at doi:10.5281/zenodo.23990216^29^.

### Contributors

HF conceived and designed the study, collected and analysed the data, coded the validation samples, and wrote the manuscript. HF is the guarantor.

### Use of artificial intelligence

Response documents were classified by a large language model (Claude Opus 5.5, Anthropic) through its batch interface, using a prompt and protocol fixed before the test set was scored. An earlier run with Gemini 2.5 Pro (Google) was scored on the same test set and superseded when the classifier was changed under a protocol amendment; its labels were not used in the analysis. Every validation label was coded by the author, blind to the model’s output. Claude Code (Anthropic) was used as a coding assistant for the collection and analysis scripts and as a drafting aid for the manuscript. The author reviewed the code and text and takes responsibility for the content; every reported number was checked by script against the analysis outputs, and every reference was checked against its source record.

