## Supplementary Materials for "Most coronial recommendations in Australia have no published response: a national analysis of findings and agency responses"

All tables were generated from the analysis outputs by the deposited code (doi:10.5281/zenodo.23990216).

Contents: Table S1; Table S2; Table S3; Table S4; Table S5; Table S6; Table S7; Table S8; Table S9; Table S10; supplementary methods; reporting checklist.

### Table S1. Linkage of response documents to findings

| Jurisdiction | Linkage key | Responses | Linked, n (%) | Unlinked |
|---|---|---|---|---|
| NSW | Deceased name | 681 | 390 (57.3) | 291 |
| Vic | Court reference (COR) | 788 | 787 (99.9) | 1 |
| Qld | Court reference, then name | 209 | 205 (98.1) | 4 |
| WA | Deceased name | 195 | 182 (93.3) | 13 |
| SA | Deceased name | 87 | 81 (93.1) | 6 |
| Tas | Court reference (TASCD) | 58 | 58 (100.0) | 0 |
| NT | Court reference (D/A number) | 22 | 22 (100.0) | 0 |

Responses with usable text (table S2 also counts empty pages). Of the 291 unlinked New South Wales responses, 207 named a person with no New South Wales finding on AustLII; 57 carried no name the linkage could use (mostly suppressed names, and inquiries into fires or groups of deaths); 22 named a person who appears in the title of a finding, mostly a joint inquest, and may be missed links; and 5 matched only findings excluded for unusable text.

### Table S2. Status of response documents by jurisdiction

| Status | NSW | Vic | Qld | WA | SA | Tas | NT | Total |
|---|---|---|---|---|---|---|---|---|
| Agency response (counts) | 589 | 778 | 209 | 194 | 87 | 55 | 22 | 1934 |
| Response exists, text unavailable (counts) | 8 | 10 | 0 | 0 | 0 | 2 | 0 | 20 |
| Court table: response awaited | 74 | 0 | 0 | 0 | 0 | 0 | 0 | 74 |
| Court table or empty page: no response recorded | 22 | 2 | 0 | 0 | 0 | 4 | 0 | 28 |
| Court table: response not required | 5 | 0 | 0 | 0 | 0 | 0 | 0 | 5 |

| Status | NSW | Vic | Qld | WA | SA | Tas | NT | Total |
|---|---|---|---|---|---|---|---|---|
| Not a response | 2 | 0 | 0 | 1 | 0 | 0 | 0 | 3 |
| Total | 700 | 790 | 209 | 195 | 87 | 61 | 22 | 2064 |

The Australian Capital Territory has no response database. Only the first two statuses count as a response; documents whose text was unavailable are excluded from the classification of acceptance.

### Table S3. Agreement with the author's blinded coding on the 100-document test set

| Classifier | Scheme | n | Accuracy (%) | Kappa (95% CI) |
|---|---|---|---|---|
| Language model (Claude Opus 5.5) | seven-category | 100 | 88.0 | 0.81 (0.71 to 0.90) |
| Language model (Claude Opus 5.5) | three-category | 100 | 92.0 | 0.79 (0.65 to 0.92) |
| Language model (Claude Opus 5.5) | binary acceptance (excl. unclassifiable) | 90 | 93.3 | 0.73 (0.50 to 0.91) |
| Rule-based (earlier) | seven-category | 100 | 32.0 | 0.17 (0.07 to 0.27) |
| Rule-based (earlier) | three-category | 100 | 49.0 | 0.22 (0.11 to 0.34) |
| Rule-based (earlier) | binary acceptance (excl. unclassifiable) | 78 | 52.6 | 0.10 (-0.01 to 0.24) |

Kappa confidence intervals from 2,000 bootstrap resamples. Binary acceptance excludes documents either rater coded unclassifiable. The test set was scored once.

### Table S4. Confusion matrix, language model against the author's coding (test set)

| Author (rows), model (columns) | implemented | already implemented | partially accepted | under consideration | noted | not supported | unclassifiable |
|---|---|---|---|---|---|---|---|
| implemented | 53 | 2 | 2 | 2 | 1 | 0 | 0 |
| already implemented | 0 | 3 | 0 | 0 | 0 | 0 | 0 |
| partially accepted | 0 | 0 | 14 | 0 | 0 | 3 | 0 |
| under consideration | 0 | 0 | 0 | 4 | 0 | 0 | 0 |
| noted | 0 | 0 | 0 | 0 | 1 | 0 | 1 |
| not supported | 0 | 0 | 0 | 1 | 0 | 4 | 0 |
| unclassifiable | 0 | 0 | 0 | 0 | 0 | 0 | 9 |

### Table S5. Blinded acceptance check on the full corpus

| Model code | Responses in stratum | Sampled | Author agreed, n (%; 95% CI) |
|---|---|---|---|
| Not accepted | 301 | 30 | 25 (83; 65 to 94) |
| Accepted | 1495 | 10 | 10 (100; 69 to 100) |

Stratified random sample drawn under a rule written before sampling; Clopper-Pearson intervals. Weighted binary accuracy 97.2%; accepted share 83.2% by the model and an estimated 86.0% by the author's coding.

### Table S6. Adjusted model of acceptance, with sensitivity analyses (n = 1,541)

| Term | Primary (Firth), OR (95% CI) | Excluding NT (ML) | Any response accepted | All responses accepted | Narrow facility flags |
|---|---|---|---|---|---|
| Northern Territory | 3.08 (0.37 to 401.56) | | 3.03 (0.36 to 395.57) | 4.85 (0.61 to 628.87) | 3.07 (0.37 to 401.00) |
| Queensland | 0.63 (0.31 to 1.29) | 0.63 (0.31 to 1.29) | 0.63 (0.31 to 1.29) | 1.07 (0.56 to 2.07) | 0.64 (0.31 to 1.31) |
| South Australia | 0.89 (0.32 to 2.98) | 0.97 (0.31 to 3.03) | 0.88 (0.32 to 2.96) | 1.60 (0.61 to 5.23) | 0.91 (0.33 to 3.04) |
| Tasmania | 0.78 (0.28 to 2.66) | 0.84 (0.26 to 2.69) | 0.79 (0.28 to 2.71) | 1.27 (0.47 to 4.23) | 0.80 (0.28 to 2.74) |
| Victoria | 0.29 (0.16 to 0.50) | 0.28 (0.16 to 0.50) | 0.30 (0.16 to 0.50) | 0.51 (0.31 to 0.82) | 0.30 (0.17 to 0.51) |
| Western Australia | 0.12 (0.06 to 0.21) | 0.11 (0.06 to 0.21) | 0.14 (0.07 to 0.26) | 0.09 (0.05 to 0.16) | 0.12 (0.06 to 0.21) |
| Year (per year) | 0.98 (0.94 to 1.03) | 0.98 (0.94 to 1.03) | 0.98 (0.94 to 1.02) | 0.99 (0.95 to 1.04) | 0.98 (0.94 to 1.02) |
| Log document length | 1.03 (0.86 to 1.23) | 1.03 (0.86 to 1.24) | 1.03 (0.86 to 1.24) | 1.02 (0.85 to 1.22) | 1.04 (0.86 to 1.24) |
| Mentions Aboriginal or Torres Strait Islander identity | 0.78 (0.46 to 1.37) | 0.79 (0.45 to 1.36) | 0.81 (0.47 to 1.44) | 0.83 (0.50 to 1.41) | 0.80 (0.47 to 1.39) |
| Mentions opioids | 0.89 (0.62 to 1.29) | 0.89 (0.62 to 1.29) | 0.93 (0.64 to 1.34) | 0.86 (0.61 to 1.22) | 0.87 (0.61 to 1.26) |
| Mentions psychiatric medicines | 0.70 (0.49 to 1.01) | 0.70 (0.49 to 1.01) | 0.70 (0.49 to 1.00) | 0.69 (0.49 to 0.99) | 0.75 (0.53 to 1.08) |
| Mentions a psychiatric unit | 2.02 (1.25 to 3.35) | 2.06 (1.26 to 3.38) | 2.11 (1.30 to 3.52) | 1.61 (1.03 to 2.54) | 1.81 (1.11 to 3.05) |
| Mentions a prison or correctional setting | 1.78 (1.12 to 2.92) | 1.82 (1.12 to 2.95) | 1.83 (1.14 to 3.04) | 1.59 (1.03 to 2.50) | 1.71 (1.07 to 2.82) |
| Mentions police custody | 0.95 (0.67 to 1.38) | 0.96 (0.66 to 1.38) | 0.95 (0.66 to 1.38) | 1.14 (0.80 to 1.64) | 0.99 (0.69 to 1.43) |

Reference jurisdiction New South Wales. Primary model: Firth penalised logistic regression with profile-likelihood intervals, majority rule for findings with several responses. Excluding NT: ordinary maximum likelihood without the Northern Territory, where every finding was accepted. Narrow facility flags (post hoc): the psychiatric facility flag without the crisis-team terms and the prison flag without watch house and remand (table S7).

### Table S7. Search terms for case characteristics

| Flag | Terms (case-insensitive, whole words) |
|---|---|
| Mentions Aboriginal or Torres Strait Islander identity | aboriginal; torres strait islander; indigenous australian; first nations; ATSI; NAAJA; ATSILS; ALS service or ALS solicitor; ACCHS; aboriginal (legal or medical or health); aboriginal (community or housing or land) council; remote community; petrol sniffing; death(s) in custody royal commission; customary law; sorry business |
| Mentions opioids | heroin; fentanyl; methadone; oxycodone; codeine; tramadol; buprenorphine; naloxone; morphine; oxycontin; suboxone; opioid; opiate |
| Mentions psychiatric medicines | antipsychotic; antidepressant; benzodiazepine; olanzapine; quetiapine; risperidone; clozapine; haloperidol; aripiprazole; sertraline; fluoxetine; citalopram; escitalopram; venlafaxine; mirtazapine; amitriptyline; diazepam; alprazolam; temazepam; oxazepam; clonazepam; lorazepam; midazolam; lithium; valproate; carbamazepine |

| Flag | Terms (case-insensitive, whole words) |
|---|---|
| Mentions a psychiatric unit | psychiatric (unit or ward or facility or hospital or inpatient); mental health (unit or ward or facility or inpatient); CAT team; crisis assessment; acute (psychiatric or mental health) |
| Mentions a prison or correctional setting | prison; correctional (centre or center or facility); detention centre; remand; watch house; inmate; youth detention; youth justice centre; juvenile (detention or justice) |
| Mentions police custody | police custody; police cell(s); police station; lock-up; custody (at or in) [the] [name] police |

A finding is flagged when its text contains any listed term. The Aboriginal and Torres Strait Islander flag combines identity terms, organisation names and contextual terms. The flags mark what a finding mentions, not the place of death or a coded attribute of the deceased.

### Table S8. Benjamini-Hochberg correction across all 41 tests

| Test | p | Adjusted p | Significant after correction |
|---|---:|---:|---|
| Indigenous to Rec rate | 5e-09 | 1.5e-08 | Yes |
| Indigenous to Acceptance | 0.38 | 0.44 | No |
| COVID pre vs acute to Rec rate | 0.0032 | 0.007 | Yes |
| Toxicology to Rec rate | 2.7e-15 | 1e-14 | Yes |
| Opioid to Rec rate | 0.0083 | 0.016 | Yes |
| Psych meds to Rec rate | 2.2e-05 | 5.6e-05 | Yes |
| Med errors to Rec rate | 4.1e-12 | 1.4e-11 | Yes |
| Polypharmacy to Rec rate | 4.1e-06 | 1.1e-05 | Yes |
| Hospital setting to Rec rate | 0.21 | 0.27 | No |
| Psychiatric fac to Rec rate | 4.3e-09 | 1.4e-08 | Yes |
| Prison to Rec rate | 0.0037 | 0.0072 | Yes |
| Aged care to Rec rate | 0.63 | 0.68 | No |
| Police custody to Rec rate | 0.00058 | 0.0014 | Yes |
| Toxicology to Acceptance | 0.014 | 0.022 | Yes |
| Prison to Acceptance | 0.00086 | 0.002 | Yes |
| Psych fac to Acceptance | 0.013 | 0.022 | Yes |
| Police custody to Acceptance | 0.35 | 0.42 | No |
| Text length: Indigenous | 1.1e-105 | 2.3e-104 | Yes |
| Text length: Opioid | 6.7e-58 | 3.9e-57 | Yes |
| Text length: Psych meds | 1.3e-78 | 1.3e-77 | Yes |
| Text length: Med errors | 2.1e-66 | 1.4e-65 | Yes |
| Text length: Polypharmacy | 2.4e-35 | 1.1e-34 | Yes |
| Text length: Hospital setting | 5.1e-21 | 2.1e-20 | Yes |
| Text length: Psychiatric fac | 1e-89 | 1.4e-88 | Yes |
| Text length: Prison | 3.3e-70 | 2.7e-69 | Yes |
| Text length: Aged care | 0.011 | 0.019 | Yes |
| Text length: Police custody | 4.9e-170 | 2e-168 | Yes |
| Text length: COVID mention | 4e-42 | 2e-41 | Yes |
| M2 coeff: is_indigenous | 0.017 | 0.025 | Yes |
| M2 coeff: med_opioid | 0.017 | 0.025 | Yes |
| M2 coeff: med_psychiatric | 0.7 | 0.72 | No |
| M2 coeff: med_error | 0.02 | 0.029 | Yes |

| Test | p | Adjusted p | Significant after correction |
|---|---|---|---|
| M2 coeff: fac_prison | 0.34 | 0.42 | No |
| M2 coeff: fac_psychiatric | 0.65 | 0.68 | No |
| M2 coeff: fac_police_custody | 0.063 | 0.084 | No |
| M4 coeff: is_indigenous | 0.38 | 0.44 | No |
| M4 coeff: med_opioid | 0.54 | 0.59 | No |
| M4 coeff: med_psychiatric | 0.058 | 0.079 | No |
| M4 coeff: fac_prison | 0.014 | 0.022 | Yes |
| M4 coeff: fac_psychiatric | 0.0036 | 0.0072 | Yes |
| M4 coeff: fac_police_custody | 0.8 | 0.8 | No |

29 of 41 tests remain significant at a false discovery rate of 5%. The family includes exploratory tests of recommendation presence and document length that are not reported in the main text.

### Table S9. Findings with recommendations that have a response, under three definitions

| Jurisdiction | With recommendations | Agency response, n (%) | Awaited only, n (%) | Any linked document, n (%) | Within database years, n/N (%; 95% CI) |
|---|---|---|---|---|---|
| NSW | 435 | 206 (47.4) | 31 (7.1) | 240 (55.2) | 172/343 (50.1; 44.9 to 55.4) |
| Vic | 1664 | 700 (42.1) | 0 (0.0) | 700 (42.1) | 700/1399 (50.0; 47.4 to 52.7) |
| Qld | 565 | 161 (28.5) | 0 (0.0) | 161 (28.5) | 158/450 (35.1; 30.8 to 39.6) |
| WA | 204 | 111 (54.4) | 0 (0.0) | 112 (54.9) | 99/169 (58.6; 51.0 to 65.7) |
| SA | 620 | 73 (11.8) | 0 (0.0) | 73 (11.8) | 69/454 (15.2; 12.2 to 18.8) |
| Tas | 733 | 37 (5.0) | 0 (0.0) | 39 (5.3) | 32/549 (5.8; 4.2 to 8.1) |
| NT | 217 | 21 (9.7) | 0 (0.0) | 21 (9.7) | 12/28 (42.9; 26.5 to 60.9) |
| ACT | 42 | 0 (0.0) | 0 (0.0) | 0 (0.0) | |

Primary definition: at least one counting response (table S2). Sensitivity definition: any linked document, including court tables recording that a response was awaited. The within-database column restricts findings to the jurisdiction's response-database years up to 2023 (co-primary measure; table S10).

### Table S10. Counting response documents by jurisdiction and publication year

| Publication year | NSW | Vic | Qld | WA | SA | Tas | NT |
|---|---|---|---|---|---|---|---|
| 2005 | 0 | 0 | 0 | 0 | 1 | 0 | 0 |
| 2006 | 0 | 0 | 0 | 0 | 5 | 0 | 0 |
| 2007 | 0 | 0 | 0 | 0 | 5 | 0 | 0 |
| 2008 | 1 | 0 | 1 | 0 | 7 | 0 | 0 |
| 2009 | 29 | 4 | 5 | 0 | 4 | 0 | 0 |
| 2010 | 52 | 40 | 3 | 0 | 5 | 0 | 0 |

| Publication year | NSW | Vic | Qld | WA | SA | Tas | NT |
|---|---|---|---|---|---|---|---|
| 2011 | 58 | 50 | 7 | 0 | 7 | 0 | 0 |
| 2012 | 45 | 86 | 13 | 0 | 2 | 0 | 0 |
| 2013 | 23 | 76 | 24 | 2 | 6 | 0 | 0 |
| 2014 | 27 | 82 | 25 | 11 | 1 | 0 | 0 |
| 2015 | 36 | 84 | 26 | 29 | 4 | 7 | 0 |
| 2016 | 26 | 65 | 10 | 16 | 1 | 9 | 0 |
| 2017 | 24 | 52 | 21 | 27 | 3 | 9 | 0 |
| 2018 | 42 | 62 | 23 | 14 | 3 | 3 | 0 |
| 2019 | 38 | 56 | 9 | 26 | 2 | 3 | 3 |
| 2020 | 26 | 90 | 10 | 13 | 5 | 0 | 2 |
| 2021 | 35 | 41 | 9 | 16 | 5 | 6 | 2 |
| 2022 | 27 | 0 | 8 | 9 | 8 | 3 | 2 |
| 2023 | 43 | 0 | 11 | 13 | 9 | 12 | 5 |
| 2024 | 46 | 0 | 3 | 17 | 4 | 4 | 4 |
| 2025 | 19 | 0 | 1 | 1 | 0 | 1 | 4 |
| Total | 597 | 788 | 209 | 194 | 87 | 57 | 22 |

The response-database years in table 1 run from the first to the last year shown for each jurisdiction. The Australian Capital Territory has no response database.

### Supplementary methods

**Text extraction audit.** Every PDF in the corpus was opened page by page to count pages with a text layer, image-only pages and blank pages. Documents in which the original extraction had read only the text layer of a mixed document were re-extracted page by page, with optical character recognition for image pages.

**Victorian index pages.** AustLII stores many Victorian responses as an index page listing the agencies that responded, followed by the scanned agency letters. Where the letters were not part of the AustLII record, the listed responses were sought on the Coroners Court of Victoria website and in Internet Archive captures of it. A retrieved document was accepted only when its text named the deceased. Ambiguous candidates were decided by the author case by case, and the decisions are recorded in the code repository.

**Response-database years.** For each jurisdiction, the response database's years run from the first to the last publication year of a counting response (table S10). The within-database response rate counts findings with recommendations from those years, truncated at 2023. Publication year is a proxy for when responses were collected; a response published early in a database's first year may answer a finding from the year before.

**Classifier.** The prompt, the protocol with its dated amendments, the model codes and the author's blinded codes are deposited with the code (doi:10.5281/zenodo.23990216).

### Reporting checklist (STROBE, cross-sectional, with RECORD items)

Reporting follows the STROBE statement for cross-sectional studies (von Elm E, Altman DG, Egger M, Pocock SJ, Gøtzsche PC, Vandenbroucke JP. Lancet. 2007;370(9596):1453-7. doi:10.1016/S0140-6736(07)61602-X) and the RECORD extension for routinely collected data (Benchimol EI, Smeeth L, Guttmann A, Harron K, Moher D, Petersen I, et al. PLoS Med. 2015;12(10):e1001885. doi:10.1371/journal.pmed.1001885).

| Item | Recommendation | Where reported |
|---|---|---|
| 1 (a, b); RECORD 1.1-1.3 | Design, data source, region, time frame and linkage in title and abstract | Title; abstract Methods |
| 2 | Scientific background and rationale | Introduction, paragraphs 1-3 |
| 3 | Objectives | Introduction, final paragraph |
| 4 | Study design | Methods, data sources |
| 5 | Setting, locations, periods | Methods, data sources; table 1 (years and response-database years) |
| 6; RECORD 6.1-6.3 | Eligibility, selection from the database, validation of selection, linkage | Methods, data sources and linking; tables S1 and S2 |
| 7; RECORD 7.1 | Outcomes, exposures and the algorithms that define them | Methods, classifying responses and case characteristics; table S7 |
| 8 | Data sources and measurement | Methods; supplementary methods |
| 9 | Bias | Methods (corpus enrichment); Discussion, strengths and limitations |
| 10 | Study size | Results; table 1 |
| 11 | Quantitative variables | Methods, statistical analysis |
| 12 (a-e); RECORD 12.1-12.3 | Statistical methods, subgroups, missing data, sensitivity analyses; data access, cleaning and linkage | Methods; tables S1, S2, S6, S8 and S9; supplementary methods |
| 13; RECORD 13.1 | Numbers at each stage, selection of the study population | Methods, data sources; Results; tables S1 and S2 |
| 14 | Descriptive data | Results; table 1 |
| 15 | Outcome data | Results; table 1; figure 1 |
| 16 | Main results, unadjusted and adjusted estimates | Results; figure 2; table S6 |
| 17 | Other analyses | Results; tables S6, S8, S9 and S10 |
| 18 | Key results | Discussion, first paragraph |
| 19; RECORD 19.1 | Limitations, including those of routinely collected data | Discussion, strengths and limitations |
| 20 | Interpretation | Discussion |
| 21 | Generalisability | Discussion |
| 22; RECORD 22.1 | Funding; access to protocol, code and data | Declarations; data availability statement |
